# Splanchnic Vein Doppler After LVAD Implantation: A Bedside Tool for Guiding Decongestion and Organ Recovery

**DOI:** 10.1101/2025.08.13.25333635

**Authors:** Michael Antonopoulos, Dimitris Elaiopoulos, Michael Bonios, Ioannis Vlahodimitris, Kiriaki Kolovou, Theodora Soulele, Giorgos Konstantinou, Kiriakos Trigkidis, Paris Vlachos, Stavros Drakos, Themistocles Chamogeorgakis, Stavros Dimopoulos, VEXUS-AKI-Onassis group collaborators

## Abstract

**Background:** Noninvasive, physiology-based methods to monitor systemic venous congestion and guide volume management early after durable LVAD implantation are lacking. We investigated whether changes in portal, hepatic, and renal vein Doppler flow patterns correlate with net fluid balance and whether these changes are associated with renal function and right ventricular (RV) performance in this setting.

**Methods:** In this prospective, proof-of-concept observational study at a national referral center for mechanical circulatory support, we enrolled 20 adult patients undergoing durable LVAD implantation between June 2024 and May 2025. Each patient underwent splanchnic venous Doppler ultrasound (hepatic, portal, and renal veins) and focused echocardiography at two time points: early after ICU admission (T0) and within 7 days (T1), post-implantation. Clinical, biochemical, and device parameters were recorded at each time point. Doppler findings were available to treating physicians and could inform real-time decisions; however, no predefined Doppler-guided protocol was applied.

**Results:** Patients experienced a significant negative fluid balance (median Δ=-6034 mL; IQR:– 8145 to –3476), which was accompanied by consistent improvement in venous Doppler profiles and a significant reduction in VExUS score (p < 0.001). Changes in portal and renal vein Doppler patterns correlated with net fluid balance (rho=0.45, p=0.046 and rho=0.68, p=0.001, respectively). A trend was also observed between VExUS change and fluid balance (rho=0.45, p=0.053). Renal function improved significantly, with an increase in eGFR (p=0.033), and right ventricular function showed parallel recovery: RV systolic velocity (S’) increased (p=0.001), RV dilation regressed (p=0.008), and tricuspid regurgitation grade decreased (p=0.001).

**Conclusions:** Point-of-care Doppler assessment of splanchnic venous flow is feasible early after LVAD implantation and reflects dynamic changes in fluid balance, renal function, and RV performance. These findings suggest that venous Doppler ultrasound may provide a physiologic, bedside method to guide decongestive therapy in this complex population, warranting validation in larger, protocol-driven studies.

**What Is New?:**

- First prospective proof-of-concept study to evaluate hepatic, portal, and renal vein Doppler flow patterns in the early period after durable LVAD implantation.
- Demonstrates that portal and renal vein Doppler changes correlate with net fluid balance and parallel improvements in renal function and right ventricular performance.
- Shows that bedside splanchnic Doppler assessment is feasible and captures physiologic changes in systemic venous congestion not reflected by conventional parameters.

**Clinical Implications:**

- Splanchnic vein Doppler assessment provides a noninvasive, physiology-based method to track systemic venous congestion after LVAD implantation and may yield insights into organ recovery.
- These data provide a foundation for larger studies to determine whether integrating splanchnic Doppler into ICU monitoring improves post-LVAD outcomes.

## Introduction

Left ventricular assist devices (LVADs) are an established and durable therapy for selected patients with advanced heart failure, offering improved survival and quality of life. They are employed as a bridge to transplantation, lifetime destination therapy, or as a bridge to recovery in select cases [1].

Despite advances in the field, early postoperative management remains challenging, particularly with respect to fluid balance and organ support. In the immediate post-LVAD period, patients often face a complex interplay of right ventricular (RV) dysfunction, dysregulated vascular tone, systemic inflammation, and neurohormonal activation, leading to venous congestion and secondary organ injury [2,3]. The superimposed acute kidney injury (AKI), with sodium and fluid retention, frequently complicates the ICU course [4].

Venous congestion is often under-recognized in ICU patients, even with advanced hemodynamic monitoring, and represents a major contributor to adverse outcomes [5]. Traditional surrogates—such as central venous pressure (CVP) and inferior vena cava (IVC) evaluation—have shown limited accuracy in predicting volume status or guiding fluid removal [6,7]. Thus, reliable, real-time tools to quantify systemic congestion and tailor decongestive strategies are critically needed.

Point-of-care ultrasound (POCUS) offers a physiologic, noninvasive bedside tool for volume assessment [8]. The Venous Excess Ultrasound Score (VExUS)—which integrates inferior vena cava assessment with Doppler interrogation of hepatic, portal, and renal veins—has emerged as a physiologically grounded tool for evaluating venous congestion. Although validated in general ICU and cardiac surgery populations [9,10], its applicability in LVAD patients—given their distinct physiology—has not been studied.

In this prospective observational study, we evaluated whether serial splanchnic Doppler changes reflect clinical decongestion in patients early after LVAD implantation. Our primary objective was to assess the correlation of vein Doppler flow changes with net fluid balance. Secondary objectives included exploring associations with renal function and RV performance.

## Methods

### Study Design, Setting and Ethics Declaration

This prospective observational study was conducted in the Cardiothoracic Surgery Intensive Care Unit of the Onassis Cardiac Surgery Center, the national referral center for heart and lung transplantation, durable mechanical circulatory support and the only active ELSO center in Greece. Informed consent was obtained from all patients (or their legal representative).The study was conducted according to the declaration of Helsinki and approved by the hospital’s research ethics committee (approval no. 809/06.06.2024).

### Patient Population

All adult patients admitted to the ICU after implantation of a durable continuous-flow LVAD between June 2024 and May 2025 were included in the study. Exclusion criteria were defined as follows: 1) age < 18 years, 2) end-stage chronic kidney disease, defined as estimated glomerular filtration rate < 15 ml/min/1.73 m² or on regular renal replacement therapy (RRT) prior to ICU admission, 3) primary hepatic parenchymal or vascular pathology impeding the ultrasound evaluation, such as established hepatic cirrhosis or thrombosis of hepatic or portal veins.

### Ultrasound Protocol

Two serial point-of-care ultrasound (POCUS) assessments were performed: the first within 72 hours of ICU admission post-LVAD implantation (T0), and the second within seven days of the initial evaluation (T1). Each session included Doppler assessment of the splanchnic veins and focused transthoracic echocardiography (fTTE). Dedicated probes were used for abdominal ultrasound and fTTE, specifically the convex (GE HealthCare 4c-Rs) and the cardiac (GE 3Sc-RS) probe respectively. All examinations were performed by a certified cardiologist-intensivist and reviewed offline. A second member of the ICU team provided an independent blinded evaluation of the POCUS results. In cases of discrepancy, a third senior intensivist provided adjudication.

At each time point, the inferior vena cava (IVC) was first assessed for dilation (defined as diameter ≥2 cm) and reduced respiratory variability (<50% collapse with inspiration). This was followed by pulsed-wave Doppler evaluation of the hepatic, portal, and intrarenal veins. Abnormal venous flow was defined as a portal vein pulsatility index greater than 30% (mild) or 50% (severe); a hepatic vein systolic wave smaller than the diastolic wave (mild) or showing systolic flow reversal (severe); and discontinuous biphasic (mild) or monophasic (severe) intrarenal venous flow. Each vein was semi-quantitatively graded for congestion severity (0 = normal, 1 = mildly abnormal, 2 = severely abnormal). The composite Venous Excess Ultrasound Score (VExUS) was calculated according to the grading system proposed by Soliman-Aboumarie et al. [11], incorporating both IVC morphology and Doppler findings: VExUS grade 0 was assigned if the IVC diameter was <2 cm, regardless of Doppler findings; grade 1 if the IVC was ≥2 cm with only normal or mildly abnormal Doppler patterns; grade 2 if one severely abnormal Doppler pattern was present; and grade 3 if two or more severely abnormal vein patterns were present. This structured approach allowed for a consistent and physiology-based assessment of systemic venous congestion at the bedside.

The fTTE assessment was performed in the parasternal long-axis, apical four-chamber, and subxiphoid views. The parameters recorded included the right ventricular basal end-diastolic diameter (EDD), RV tissue Doppler velocity (RVS’, cm/sec), and the severity of tricuspid regurgitation (TR). The echocardiographic RV indices were collected and evaluated in accordance with the guidelines established by Lang RM et al [12]. An RV was classified as dilated if the EDD was ≥40 mm, and TR was graded qualitatively, with 0 representing no regurgitation, 1 for mild, 2 for moderate, and 3 for severe regurgitation.

### Clinical Data Collection and Definitions

Demographic variables, comorbidities, and anthropometric data were collected at baseline. Clinical characteristics (serum creatinine, serum urea, lactate, total bilirubin, NTproBNP, fluid balance, vasoactive inotropic score, central venous pressure, LVAD functional parameters) were recorded and documented both for the day of the first (T0) and second (T1) ultrasound evaluation.

The numerical difference (Δ-) for all assessed variables was determined by subtracting the values recorded on time point T0 from those on time point T1. For variables that had multiple measurements on the same day, the measurement that occurred closest to the ultrasound assessment, within a 12-hour period, was employed. Vasoactive-inotropic score (VIS) was calculated according to published guidance from Belletti et al [13]. Glomerular filtration rate (eGFR) was estimated by using the CKD-EPI creatinine 2021 formula [14].

### Integration of Doppler Findings into Clinical Care

Although no standardized protocol governed fluid management, Doppler findings were available to treating ICU physicians and informed their clinical decision-making. When markedly abnormal patterns were observed—such as monophasic intrarenal flow, portal pulsatility >50%, or a VExUS score ≥2—decongestive measures were frequently initiated, including diuretic adjustment or RRT, as appropriate. These decisions were individualized and made at the discretion of the treating team, rather than being dictated solely by ultrasound results.

## Statistical analysis

All continuous variables are reported as medians (25th-75th percentiles). Categorical variables are reported as absolute numbers (n) and percentages (%). All continuous variables were tested for normality distribution by Kolmogorov-Smirnov test. The Wilcoxon signed-rank test was used to compare paired data. Fisher’s exact test was used to determine whether there is a statistically significant difference between the expected and the observed frequencies in one or more categories of a contingency table. Correlation analyses were performed using Spearman’s rank correlation coefficient (rho). Scatterplots with fitted trend lines were used to visualize correlations between Doppler vein flow changes and other clinical variables. Statistical significance was defined as two-tailed p value < 0.05. The SPSS statistical program (v.24; SPSS, Chicago, IL) was used for data analysis.

## Results

### Patient Population

During the study period 20 patients underwent LVAD insertion and met inclusion criteria. All patients were implanted with a HeartMate3™ LVAD (Abbott, Chicago, IL, U.S.). One patient was supported with a second HM3 device configured as an RVAD due to intractable electrical storm, and another required temporary veno-pulmonary ECMO for severe right ventricular dysfunction post-implantation. The first POCUS assessment was performed at a median of 3 days post-ICU admission, and the second at a median of 4 days thereafter. Baseline characteristics and clinical outcomes are shown in Table 1.

**Table 1.** Baseline Demographics, Comorbidities, and Clinical Outcomes of the Study Population (n = 20)

|  |  |
| --- | --- |
| **Demographics** |  |
| Sex, male, n (%) | 19 (95) |
| Age, years | 49 (37 - 56) |
| BMI, kg/m<sup>2</sup> | 24.6 (21.7 - 28.6) |
| **Etiology of LVAD implantation, n (%)** |  |
| Dilated Cardiomyopathy | 14 (70) |
| Ischemic Cardiomyopathy | 5 (25) |
| Other | 1 (5) |
| **Comorbidities, n (%)** |  |
| Diabetes mellitus | 5 (25) |
| Arterial hypertension | 4 (20) |
| Atrial fibrillation | 8 (40) |
| Chronic kidney disease | 9 (45) |
| Ischemic stroke | 4 (20) |
| Peripheral arterial disease | 1 (5) |
| COPD | 1 (5) |
| Autoimmune disease | 1 (5) |
| **Clinical Outcomes** |  |
| ICU LOS, days | 11 (6-16) |
| Mechanical Ventilation, hours | 35 (14-75) |
| RRT ICU, n (%) | 2 (10) |
| Survival ICU, n (%) | 19 (95) |
| Survival to discharge, n (%) | 19 (95) |

BMI, body mass index; COPD, chronic obstructive pulmonary disease; ICU, intensive care unit; LOS, length of stay; RRT, renal replacement therapy All continuous variables are presented as median (25<sup>th</sup> -75<sup>th</sup> percentiles)

### Clinical Trends Between First and Second POCUS assessments

Patients experienced a significant median negative fluid balance between the two serial assessments, paralleled by improvements in key clinical markers. Serum creatinine decreased (median ΔCr = –0.3 mg/dL) and eGFR increased (median ΔeGFR = +12 mL/min/1.73 m²). Renal function improved in 12 patients (60%), remained stable in 3 (15%), and worsened in 5 (25%). This trend persisted until ICU discharge (median ΔeGFR= +15 mL/min/1.73 m²), which occurred at a median of 4 days after the second evaluation. CVP and NT-proBNP values declined, while lactate and total bilirubin levels improved. Changes in hemodynamic, laboratory, and device parameters between first and second POCUS assessments (T0 vs T1) are presented in Table 2.

**Table 2.**
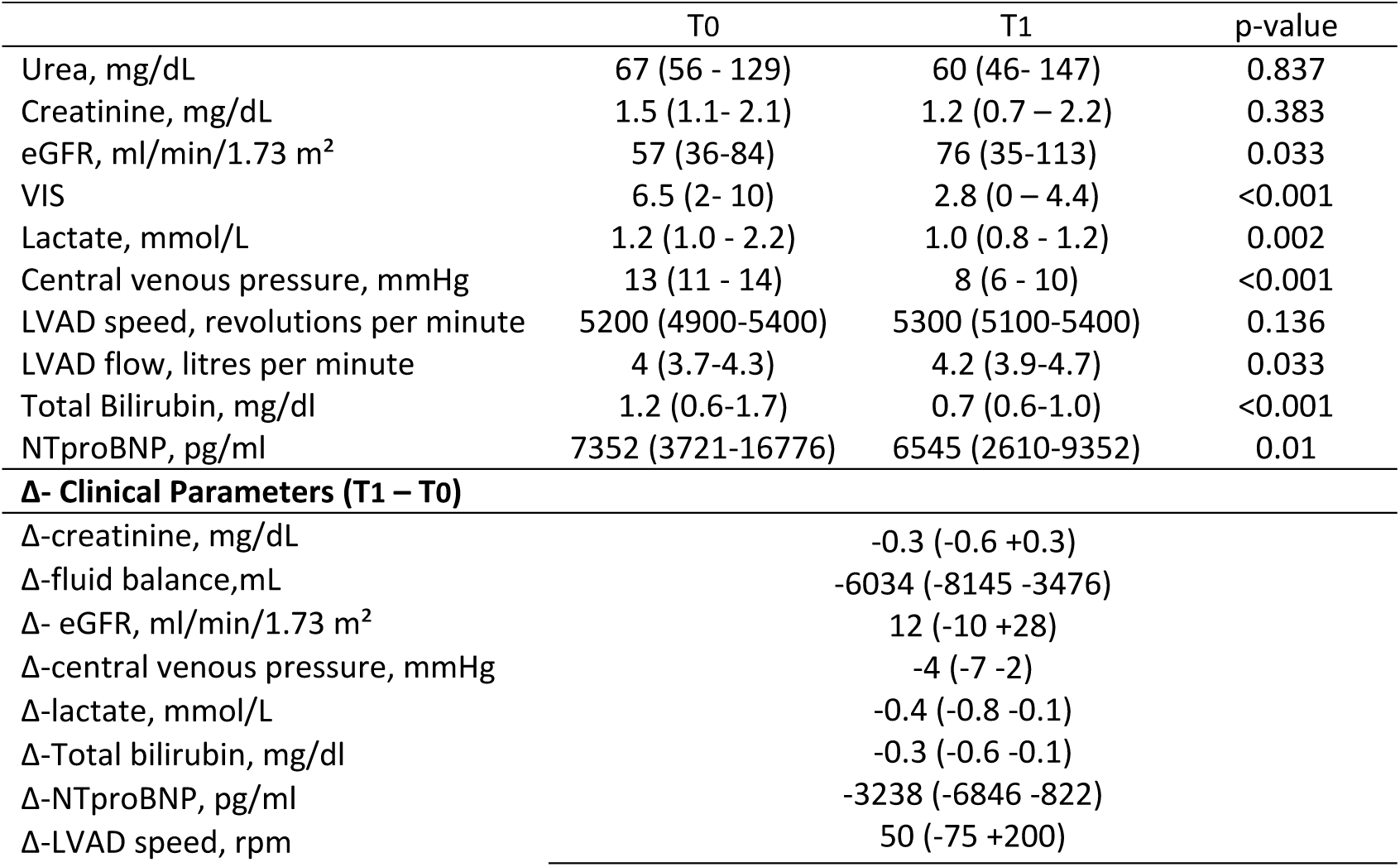
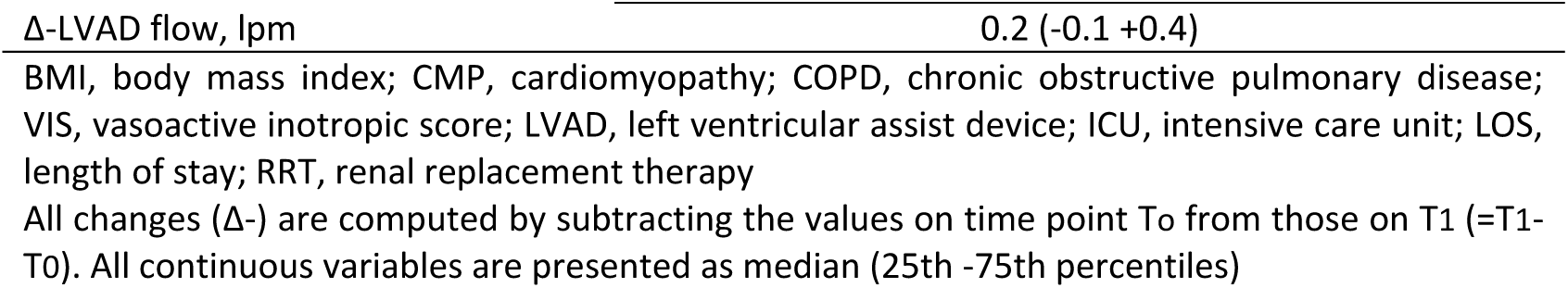
Hemodynamic, Laboratory, and Device parameter changes between first and second ultrasound assessments (T0 vs T1)

|  | T0 | T1 | p-value |
| --- | --- | --- | --- |
| Urea, mg/dL | 67 (56 - 129) | 60 (46- 147) | 0.837 |
| Creatinine, mg/dL | 1.5 (1.1- 2.1) | 1.2 (0.7 – 2.2) | 0.383 |
| eGFR, ml/min/1.73 m<sup>2</sup> | 57 (36-84) | 76 (35-113) | 0.033 |
| VIS | 6.5 (2- 10) | 2.8 (0 – 4.4) | <0.001 |
| Lactate, mmol/L | 1.2 (1.0 - 2.2) | 1.0 (0.8 - 1.2) | 0.002 |
| Central venous pressure, mmHg | 13 (11 - 14) | 8 (6 - 10) | <0.001 |
| LVAD speed, revolutions per minute | 5200 (4900-5400) | 5300 (5100-5400) | 0.136 |
| LVAD flow, litres per minute | 4 (3.7-4.3) | 4.2 (3.9-4.7) | 0.033 |
| Total Bilirubin, mg/dl | 1.2 (0.6-1.7) | 0.7 (0.6-1.0) | <0.001 |
| NTproBNP, pg/ml | 7352 (3721-16776) | 6545 (2610-9352) | 0.01 |
| **Δ- Clinical Parameters (T1 – T0)** |  |  |  |
| Δ-creatinine, mg/dL |  | -0.3 (-0.6 +0.3) |  |
| Δ-fluid balance,mL |  | -6034 (-8145 -3476) |  |
| Δ- eGFR, ml/min/1.73 m<sup>2</sup> |  | 12 (-10 +28) |  |
| Δ-central venous pressure, mmHg |  | -4 (-7 -2) |  |
| Δ-lactate, mmol/L |  | -0.4 (-0.8 -0.1) |  |
| Δ-Total bilirubin, mg/dl |  | -0.3 (-0.6 -0.1) |  |
| Δ-NTproBNP, pg/ml |  | -3238 (-6846 -822) |  |
| Δ-LVAD speed, rpm |  | 50 (-75 +200) |  |

|  |  |
| --- | --- |
| Δ-LVAD flow, lpm | 0.2 (-0.1 +0.4) |

BMI, body mass index; CMP, cardiomyopathy; COPD, chronic obstructive pulmonary disease; VIS, vasoactive inotropic score; LVAD, left ventricular assist device; ICU, intensive care unit; LOS, length of stay; RRT, renal replacement therapy
All changes ( $\Delta$ -) are computed by subtracting the values on time point T0 from those on T1 (=T1-T0). All continuous variables are presented as median (25th -75th percentiles)

### Doppler Vein Flow Assessment

All three splanchnic veins were successfully visualized and scored in all 20 patients at both time points, yielding a VExUS feasibility rate of 100%. The two consecutive ultrasonographic evaluations showed consistent trends of improved flow patterns across all the examined splanchnic veins. A case from our cohort is illustrated in figure 1. All patients demonstrated improvement in at least one Doppler component, with a median ΔVExUS of-2. Improvement in two components was observed in 10 patients (50%), and in all three in 8 patients (40%). The median ΔPortal vein was - 1, which was also true for both ΔHepatic and ΔRenal veins. VExUS score and its individual components on time points T0 and T1 are described in Table 3.

**Figure 1.**
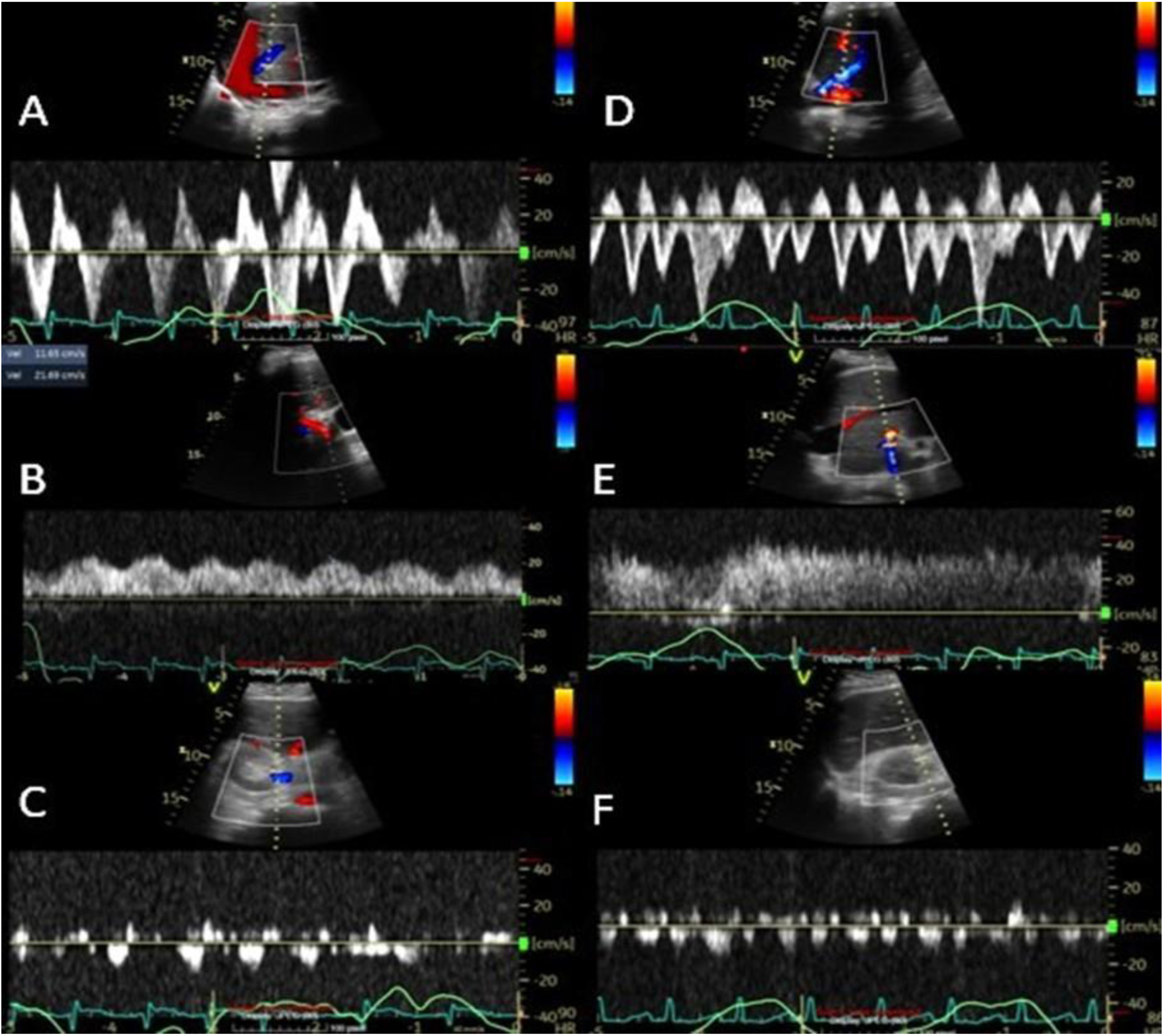
Changes in splanchnic vein Doppler profiles between first and second ultrasound assessments post LVAD implantation (T0 vs T1), in LVAD patient n 7 from our cohort. Severe venous congestion early post LVAD implantation: A) Systolic reversal of hepatic venous flow, B) Portal vein pulsatility index of 50%, C) Monophasic intrarenal venous flow. Decongestion after aggressive diuresis (fluid balance of-6400 ml): D) S<D forward flow waves in hepatic vein, E) Portal vein flow with minimal pulsatility, F) Biphasic intrarenal venous flow

**Table 3.**
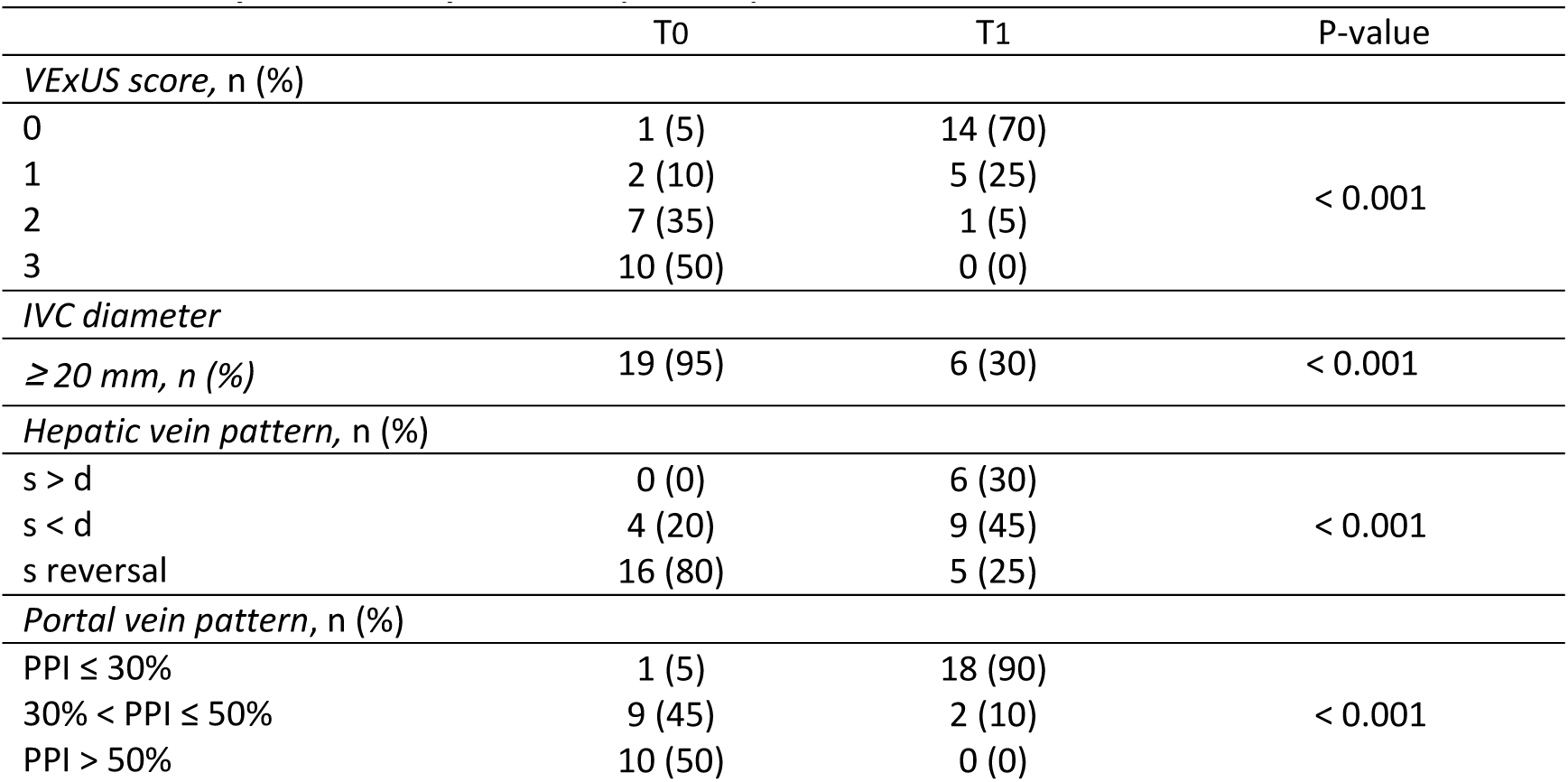
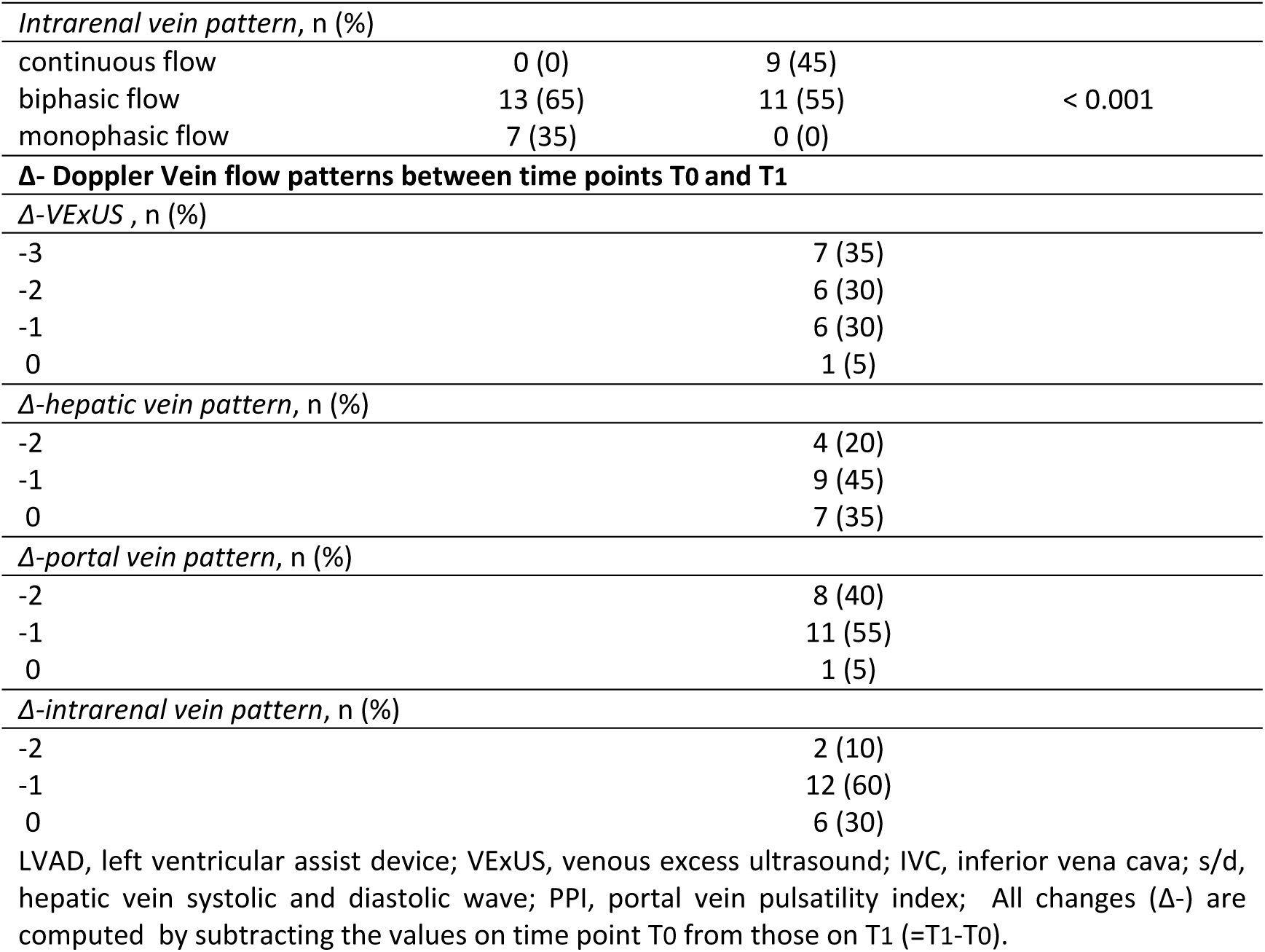
Changes in VExUS score and its components between first and second ultrasound assessments post LVAD implantation (T0 vs T1)

|  | T0 | T1 | P-value |
| --- | --- | --- | --- |
| *VExUS score, n (%)* |  |  |  |
| 0 | 1 (5) | 14 (70) | < 0.001 |
| 1 | 2 (10) | 5 (25) |  |
| 2 | 7 (35) | 1 (5) |  |
| 3 | 10 (50) | 0 (0) |  |
| *IVC diameter* |  |  |  |
| $\geq 20$ mm, n (%) | 19 (95) | 6 (30) | < 0.001 |
| *Hepatic vein pattern, n (%)* |  |  |  |
| s > d | 0 (0) | 6 (30) | < 0.001 |
| s < d | 4 (20) | 9 (45) |  |
| s reversal | 16 (80) | 5 (25) |  |
| *Portal vein pattern, n (%)* |  |  |  |
| PPI $\leq 30\%$ | 1 (5) | 18 (90) | < 0.001 |
| $30\% < \text{PPI} \leq 50\%$ | 9 (45) | 2 (10) | |
| PPI > 50% | 10 (50) | 0 (0) |  |
| *Intrarenal vein pattern, n (%)* |  |  |  |
| continuous flow | 0 (0) | 9 (45) | < 0.001 |
| biphasic flow | 13 (65) | 11 (55) |  |
| monophasic flow | 7 (35) | 0 (0) |  |
| **Δ- Doppler Vein flow patterns between time points T0 and T1** |  |  |  |
| *Δ-VExUS , n (%)* |  |  |  |
| -3 |  | 7 (35) |  |
| -2 |  | 6 (30) |  |
| -1 |  | 6 (30) |  |
| 0 |  | 1 (5) |  |
| *Δ-hepatic vein pattern, n (%)* |  |  |  |
| -2 |  | 4 (20) |  |
| -1 |  | 9 (45) |  |
| 0 |  | 7 (35) |  |
| *Δ-portal vein pattern, n (%)* |  |  |  |
| -2 |  | 8 (40) |  |
| -1 |  | 11 (55) |  |
| 0 |  | 1 (5) |  |
| *Δ-intrarenal vein pattern, n (%)* |  |  |  |
| -2 |  | 2 (10) |  |
| -1 |  | 12 (60) |  |
| 0 |  | 6 (30) |  |

LVAD, left ventricular assist device; VExUS, venous excess ultrasound; IVC, inferior vena cava; s/d, hepatic vein systolic and diastolic wave; PPI, portal vein pulsatility index; All changes (Δ-) are computed by subtracting the values on time point T0 from those on T1 (=T1-T0).

Changes in splanchnic venous Doppler flow patterns demonstrated significant correlations with net fluid balance, as illustrated in Figure 2. Among individual components, renal vein flow changes correlated strongly with net fluid balance (rho = 0.68, p = 0.001), followed by portal vein flow changes (rho = 0.45, p = 0.046), while hepatic vein changes showed a non-significant trend (rho = 0.364, p = 0.11). The composite VExUS score demonstrated a trending association (rho = 0.454, p = 0.053), with greater Doppler improvement aligning with more negative fluid balance. CVP (rho = 0.125, p = 0.599) and IVC diameter (rho = 0.059, p = 0.806) did not correlate with fluid balance.

**Figure 2.**
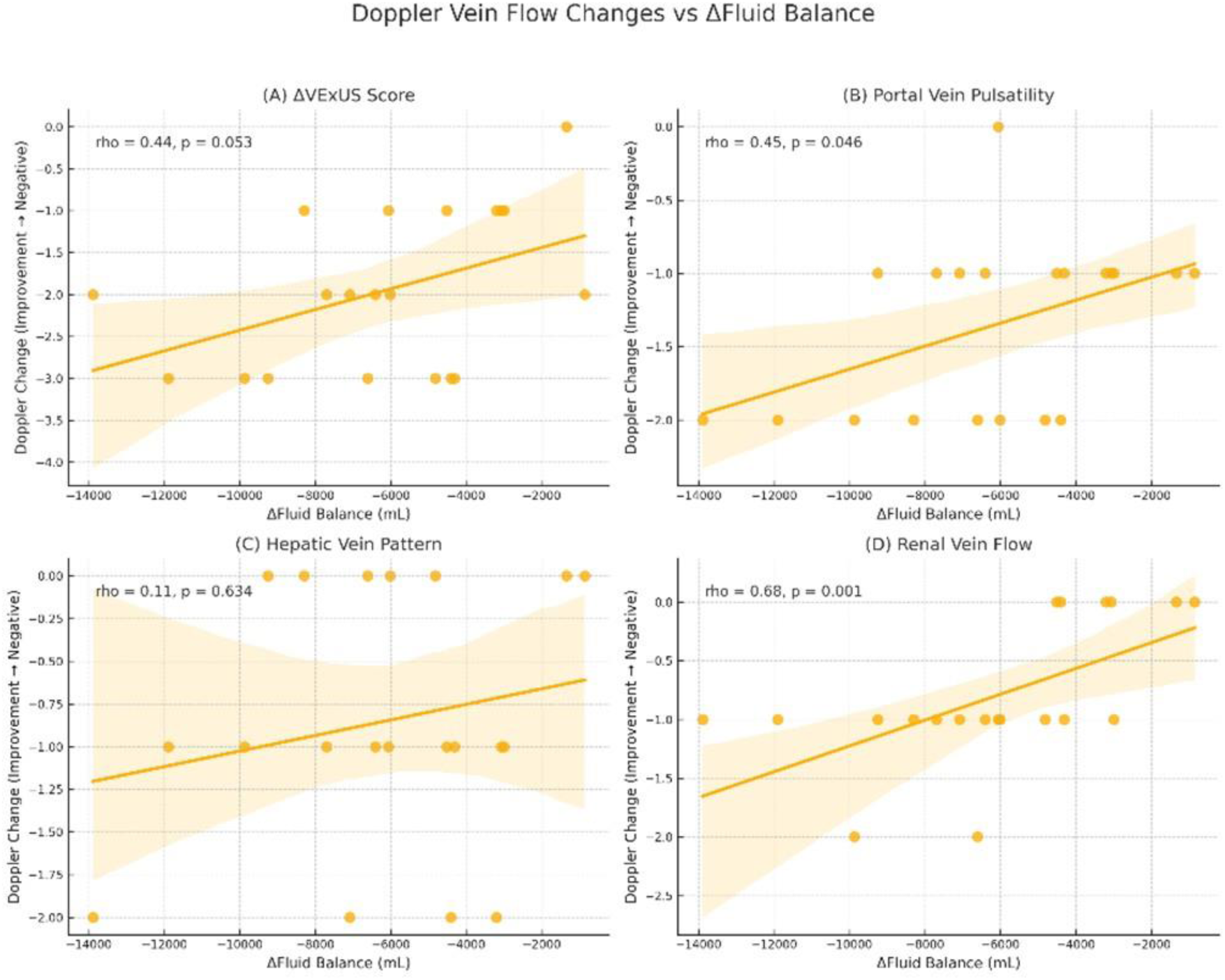
Correlation between Doppler Vein Flow Changes and ΔFluid Balance in LVAD patients over the ultrasound evaluation period. Multi-panel scatterplots showing associations between net fluid balance and changes in (A) VExUS score, (B) portal vein pulsatility, (C) hepatic vein Doppler pattern, and (D) renal vein flow. Trend lines and Spearman’s rho (ρ) with p-values are shown in each panel. Negative y-axis values reflect Doppler improvement. Findings suggest that greater decongestion is linked with favorable shifts in vein Doppler profiles, supporting the physiologic rationale for incorporating splanchnic vein Doppler as a real-time bedside tool to guide volume management in post-LVAD patients.

### Echocardiographic Findings

Right ventricular systolic function improved, as indicated by a median ΔRVS’ of +1 cm/s. RVS’ increased in 13 (65%) patients, was unchanged in 6 (30%) and declined only in one. Tricuspid regurgitation grade decreased in most patients, reflected by a median ΔTR of –1. Out of 14 patients (70%) who initially had at least moderate TR grade, only 3 (15%) retained this finding in the follow-up evaluation. RV dilation resolved in 8 of 9 affected patients. fTTE and its components on time points T0 and T1 are presented in Table 4.

**Table 4.** Focused echocardiography and its components assessed on time points T0 and T1 post LVAD implantation.

|  | T0 | T1 | p-value |
| --- | --- | --- | --- |
| RVS', cm/sec | 7 (6-8) | 8 (7-9) | 0.001 |
| RV Dilation, n (%) | 9 (45) | 1(5) | 0.008 |
| Tricuspid Regurgitation grade, n (%) |  |  | 0.001 |
| 0 | 0 (0) | 2 (10) |  |
| 1 | 6 (30) | 15 (75) |  |
| 2 | 8 (40) | 3 (15) |  |
| 3 | 6 (30) | 0 (0) |  |
| **Δ- Right ventricular echocardiographic indices between time points T0 and T1** |  |  |  |
| ΔRVS', cm/sec |  | +1 (0 -1) |  |
| ΔTR grade |  | -1 (-1 +1) |  |

RV, right ventricle; RVS', right ventricular systolic excursion velocity; TR, tricuspid regurgitation All changes (Δ-) are computed by subtracting the values on time point To from those on T1 (=T1-T0) All continuous variables are presented as median (25th -75th percentiles)

## Discussion

This proof-of-concept study demonstrates that bedside Doppler ultrasound assessment of splanchnic vein flow is feasible and may provide physiologically meaningful insights into fluid status and organ recovery early after LVAD implantation. Within the complex hemodynamic landscape of the ICU, we observed that changes in the composite Venous Excess Ultrasound Score (VExUS) and in its individual Doppler components - including hepatic, portal, and intrarenal vein flow patterns – were associated with net negative fluid balance over time. To our knowledge, this is the first prospective study to describe such dynamic associations in patients early post LVAD implantation.

Patients who experienced greater negative fluid balance over the two sequential POCUS evaluations exhibited more favorable changes in splanchnic Doppler profiles. Specifically, they demonstrated a reduction in portal vein pulsatility, a shift toward systolic-dominant hepatic vein flow, and conversion from discontinuous to continuous intrarenal venous waveforms. These findings align with prior evidence linking Doppler vein flow abnormalities to elevated right-sided pressures and systemic venous congestion in broader critical care settings [15,16,17]. As decongestion progresses — whether through diuretics or RRT — venous return pressure gradients normalize, which is mirrored in improving Doppler waveforms [18].

Our secondary findings highlight parallel improvements in renal function, right ventricular performance, and biomarkers of congestion among patients who achieved effective decongestion and favorable changes in splanchnic vein Doppler flow. The removal of excess fluid in the setting of a recovering right ventricle is thought to reduce CVP and abdominal venous congestion, thereby improving the renal perfusion pressure gradient — the difference between mean arterial and venous pressure — which is a critical determinant of glomerular filtration [19]. Indeed, elevated right atrial pressure is independently associated with impaired renal recovery and increased risk of RRT following LVAD implantation [20]. In our cohort, the rate of RRT use (10%) aligns with prior reports [21], while a trend toward sustained renal improvement was observed, up to the point of ICU discharge. Importantly, renal function recovery is associated with better outcomes in heart failure patients and LVAD recipients [22]. Even modest changes in creatinine and eGFR values (>10 ml/min/1.73 m²) may have prognostic implications [23].

Right ventricular function also improved significantly over time: RV S’ increased, RV dilation regressed, and TR grade decreased in most patients. These improvements support the hypothesis that fluid offloading may reduce RV preload and afterload, facilitate forward flow, and promote early RV reverse remodeling — a mechanism increasingly recognized in LVAD-supported patients [24]. Notably, these changes occurred despite a marked reduction in vasoactive-inotropic support (VIS), suggesting that hemodynamic optimization through volume management, rather than pharmacologic support alone, may play a dominant role in early postoperative recovery [25,26].

Importantly, our findings suggest that Doppler ultrasound may provide incremental value over traditional static markers of volume status. Neither central venous pressure (CVP) nor inferior vena cava (IVC) diameter changes correlated with fluid balance in our cohort, whereas Doppler-derived vein assessments did. This observation underscores the limitations of conventional metrics [27] and highlights the potential of splanchnic Doppler to more accurately capture clinically relevant congestion dynamics.

The improvement in CVP, alongside congestion-sensitive biomarkers such as NT-proBNP and total bilirubin, further strengthens the physiological signal that early decongestion and enhanced venous return were clinically meaningful. These findings align with previous studies highlighting the role of venous congestion in organ dysfunction and the prognostic value of biomarkers in heart failure and post-LVAD care [28,29].

Taken together, our results provide novel insights into the interplay between venous congestion, fluid management, and end-organ recovery in the early post-LVAD period. By integrating splanchnic vein Doppler into routine bedside evaluation, clinicians may gain a more granular, real-time understanding of congestion and its resolution—particularly when conventional measures fall short. While our findings are exploratory, they lay the groundwork for larger, outcome-driven studies to evaluate the utility of Doppler-guided volume strategies in this high-risk population.

## Limitations

This single-center study included a small sample size, which may limit statistical power and restrict external validity; however, this was a pilot study accompanied by statistical significant results, supporting our conclusions. The observational design precludes causal inference, and although the timing of assessments was standardized, it may not fully capture the dynamic fluid shifts and hemodynamic variability characteristic of the early post-LVAD period. VExUS is a dynamic and operator-dependent modality. While formal interobserver variability was not quantified, all ultrasound assessments were performed by experienced operators, independently double-reviewed, and adjudicated by a senior intensivist to enhance internal consistency. A previous study has already shown high reproducibility of VEXUS assessment [30]. Additionally, while consistent associations were observed between Doppler changes and net fluid balance, residual confounding from individualized medical decisions—such as diuretic dosing or right heart loading conditions—cannot be entirely excluded. Despite these limitations, the physiologic coherence of our findings supports further investigation in larger prospective studies focused on clinical outcomes and external validation.

## Conclusion

This prospective, exploratory study demonstrates that splanchnic vein Doppler ultrasound is feasible and may act as a physiologically informative tool to guide fluid management in patients early after LVAD implantation. Changes in portal, hepatic, and intrarenal venous Doppler flow patterns tracked with negative fluid balance, renal function recovery, and right ventricular improvement. These findings support the potential utility of Doppler-based congestion assessment as a real-time, noninvasive adjunct to guide decongestive therapy and monitor right heart unloading in this high-risk population.

## Non-standard Abbreviations and Acronyms

AKI: acute kidney injury
eGFR: estimated glomerular filtration rate
CKD EPI: chronic kidney disease epidemiology collaboration.
KDIGO: kidney disease: improving global outcomes
RRT: renal replacement therapy
ICU: intensive care unit
CVP: central venous pressure
RAP: right atrial pressure
IVC: inferior vena cava
POCUS: point-of-care ultrasonography
VExUS: venous excess ultrasound score
fTTE: focused transthoracic echocardiography
ELSO: extracorporeal life support organization
LVAD: left ventricular assist device
MCS: mechanical circulatory support
HM3: HeartMate 3
RV: right ventricle/ventricular
RVS′: right ventricular systolic tissue doppler velocity at tricuspid annulus
TR: tricuspid regurgitation
EDD: end diastolic diameter
VIS: vasoactive inotropic score

## Data Availability

The data that support the findings of this study are available from the corresponding author upon reasonable request.

## Acknowledgments

Antonopoulos M: Study concept and design, Analyzed the data and wrote the manuscript; Elaiopoulos D: Concept and design, revised the manuscript; Bonios M: revised the manuscript; Vlahodimitris I: revised the manuscript; Kolovou K: revised the manuscript; Soulele T: revised the manuscript; Konstantinou G: revised the manuscript; Trigkidis K: revised the manuscript; Vlachos P: revised the manuscript; Drakos S: revised the manuscript; Chamogeorgakis T: revised the manuscript; and Stavros Dimopoulos: main supervisor, designed and approved the study and revised the manuscript. All authors approved the final version of this article.

## Appendix

VEXUS-AKI-Onassis group collaborators:

Antigoni Koliopoulou, Aggeliki Gouziouta, Nektarios Kogerakis, Konstantinos Ieromonachos, Thodoris Pitsolis, Chris Kapelios, Mixalis Zervos, Konstantina Kolonia, Maria Chronaki, Eleni Tzatzaki, Paraskevi Salata, Fotios Dimitriadis, Theodosia Maragkoulia, Despina Chilidou, Chrisa Panagiotou, Sissy Stergiopoulou, Vasia Tsagkari, Evangelos Leontiadis, Stamatis Adamopoulos

---

STROBE Statement—checklist of items that should be included in reports of observational studies

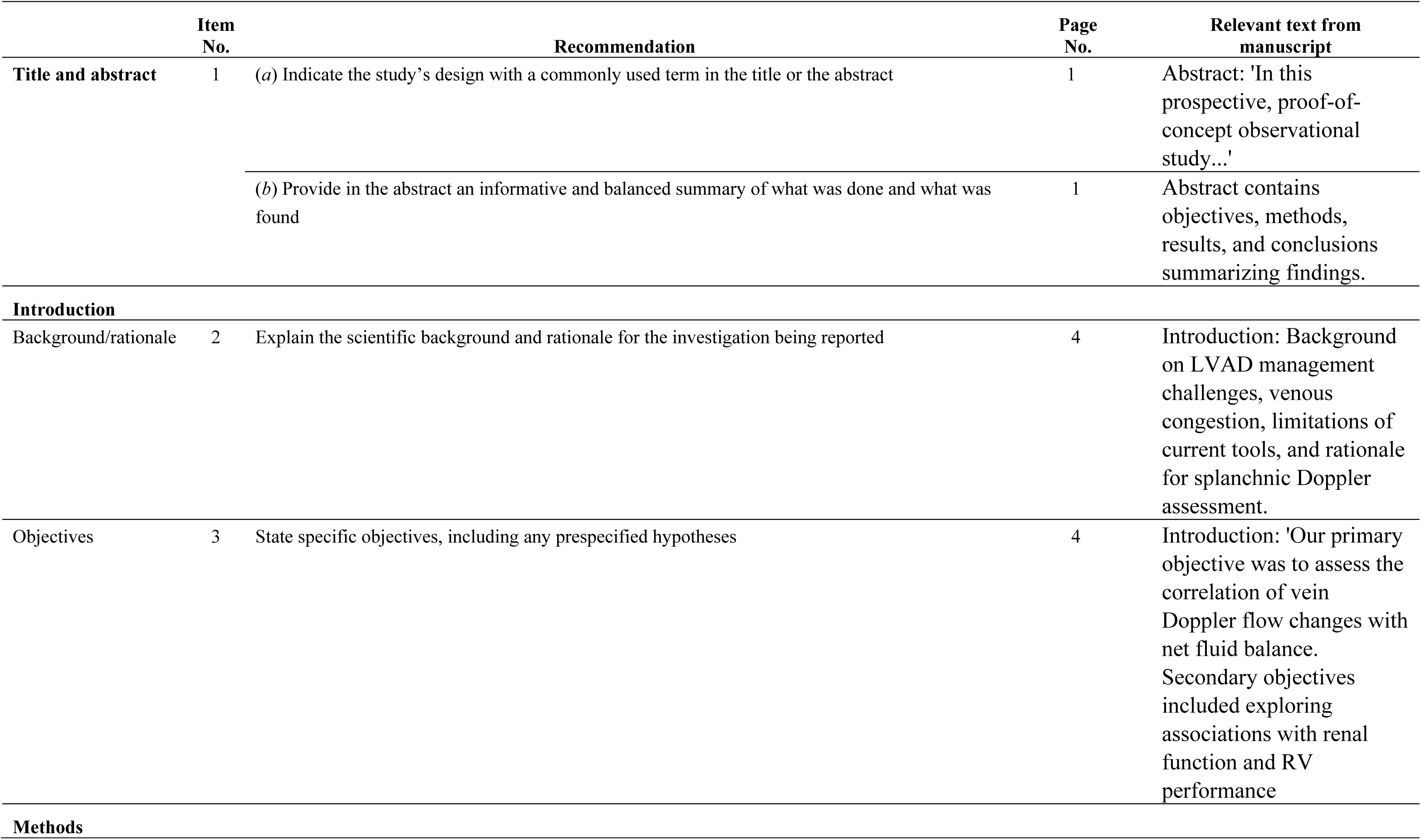
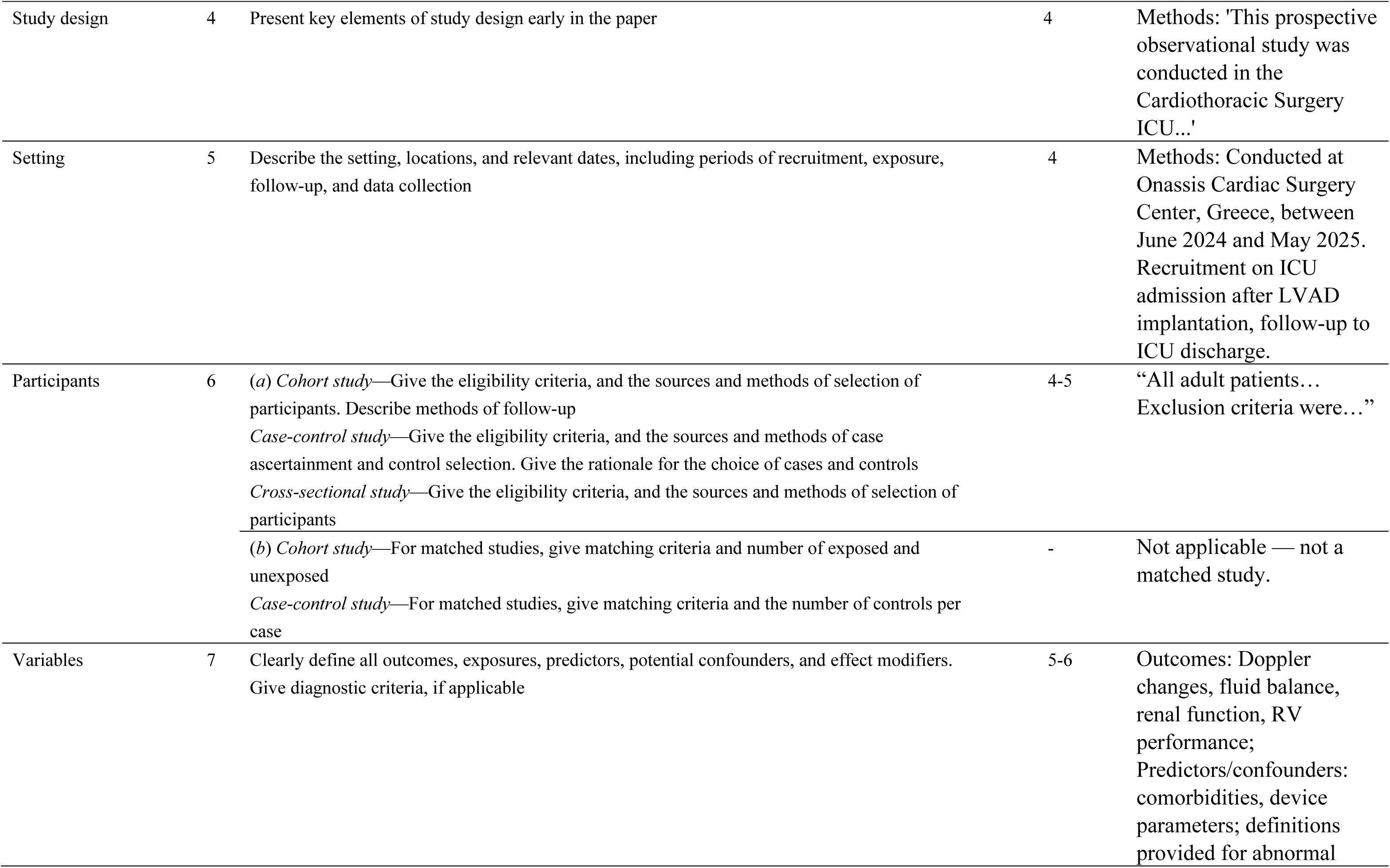
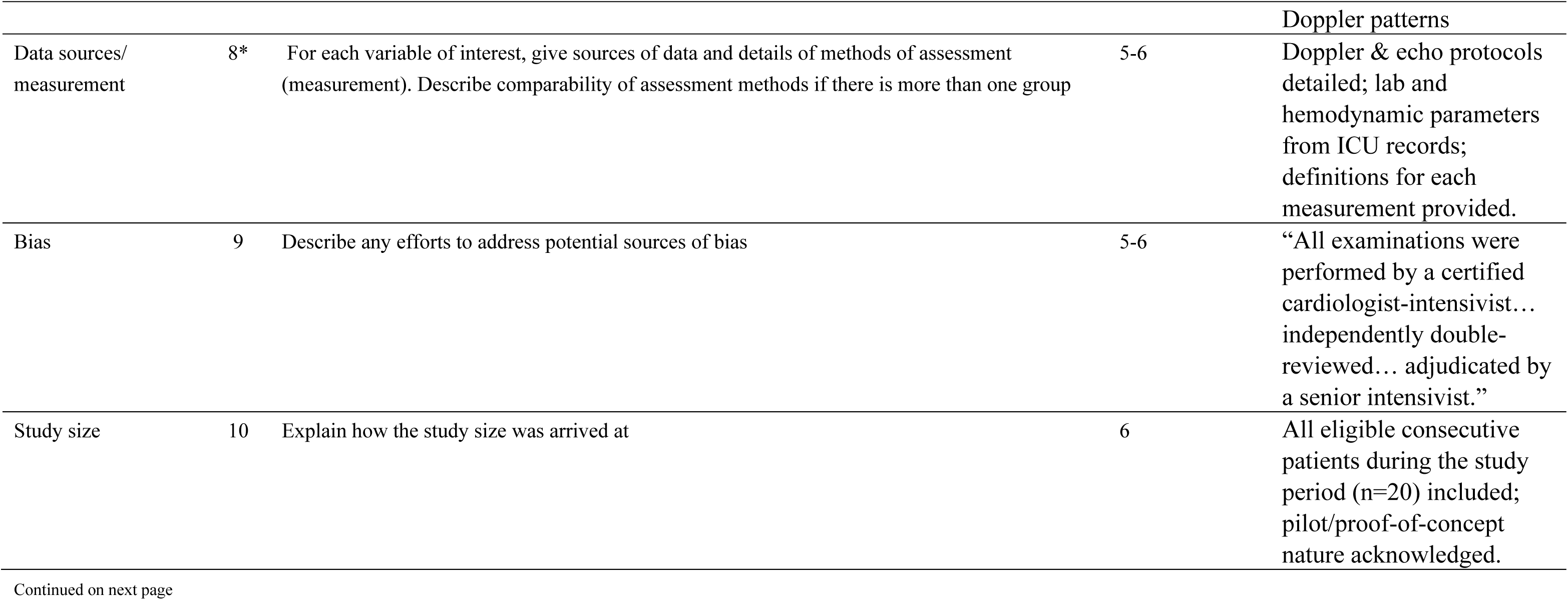
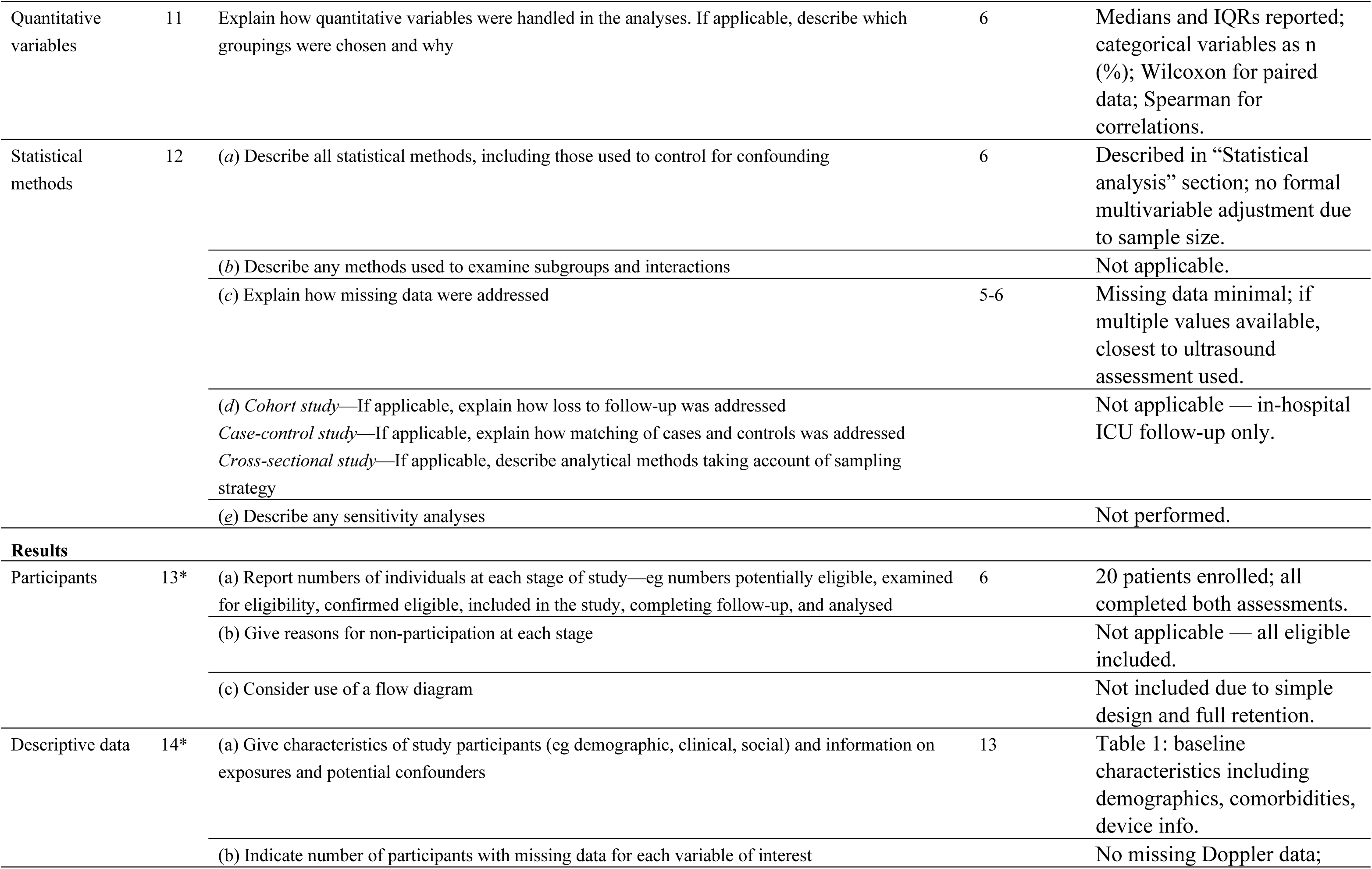
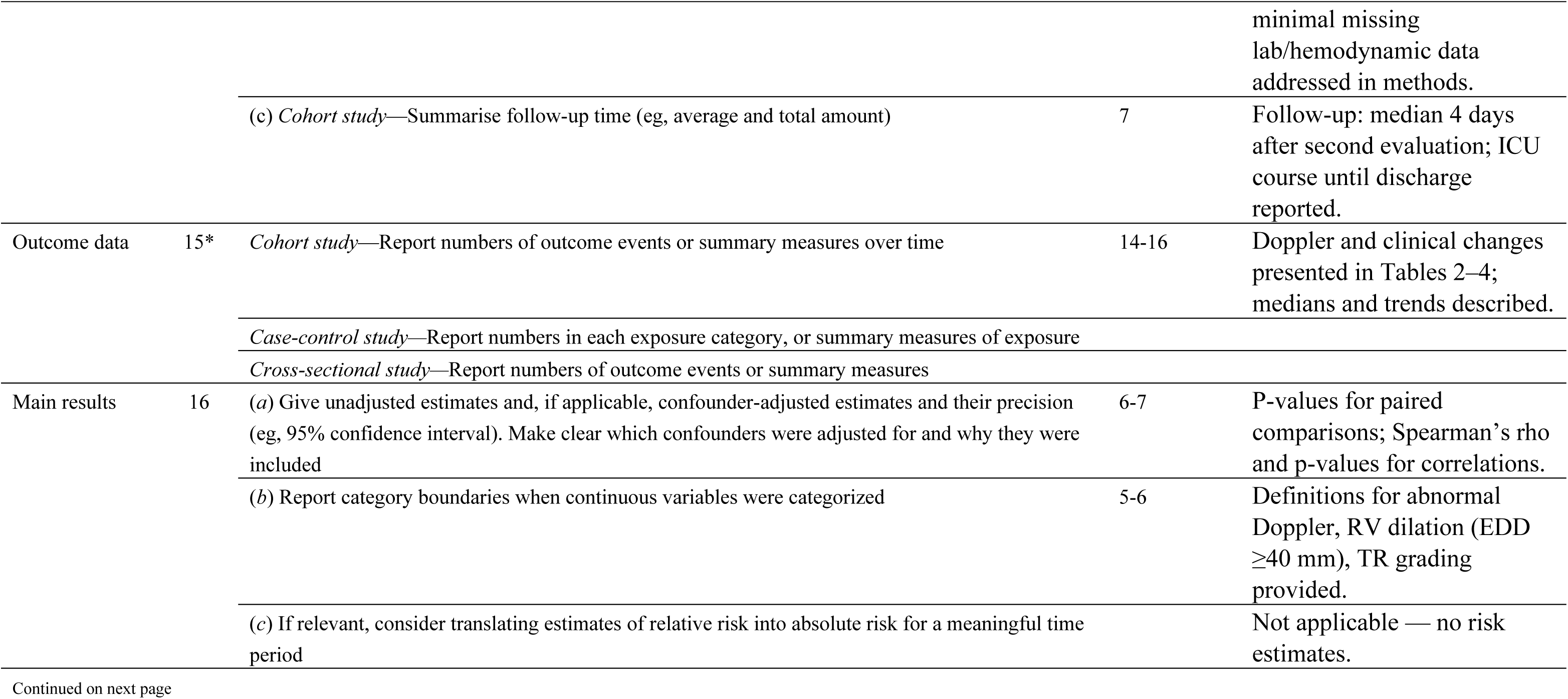
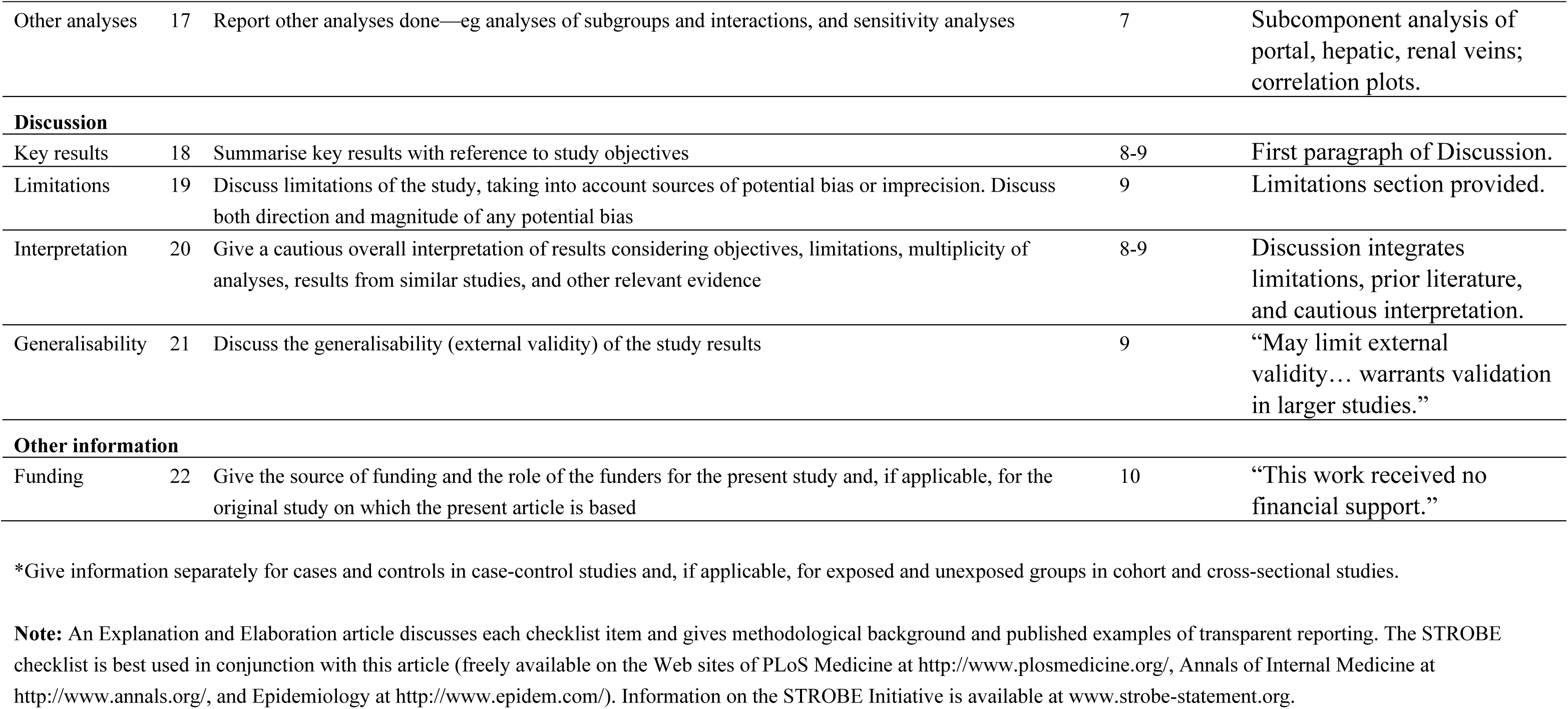

## References

1) Tedford RJ, Leacche M, Lorts A, et al. Durable Mechanical Circulatory Support: JACC Scientific Statement. J Am Coll Cardiol. 2023 Oct 3;82(14):1464–1481.

2) Walther CP, Civitello AB, Lamia HK, et al. Kidney Function Trajectories and Right Heart Failure Following LVAD Implantation. J Am Heart Assoc. 2024 Mar 5;13(5):e031305.

3) Antonopoulos M, Bonios MJ, Dimopoulos S, et al. Advanced Heart Failure: Therapeutic Options and Challenges in the Evolving Field of Left Ventricular Assist Devices. J Cardiovasc Dev Dis. 2024 Feb 16;11(2):61.

4) Rangaswami J, Bhalla V, Blair JEA, et al. Cardiorenal syndrome: classification, pathophysiology, diagnosis, and treatment strategies: a scientific statement from the American Heart Association. Circulation. 2019;139:e840–e878.

5) Kattan E, Castro R, Miralles-Aguiar F, et al. The emerging concept of fluid tolerance: A position paper. J Crit Care. 2022 Oct;71:154070.

6) Marik PE, Cavallazzi R. Does the central venous pressure predict fluid responsiveness? An updated meta-analysis and a plea for some common sense. Crit Care Med. 2013 Jul;41(7):1774–81.

7) Via G, Tavazzi G, Price S. Ten situations where inferior vena cava ultrasound may fail to accurately predict fluid responsiveness: a physiologically based point of view. Intensive Care Med. 2016 Jul;42(7):1164–7.

8) Argaiz ER, Koratala A, Reisinger N. Comprehensive Assessment of Fluid Status by Point-of-Care Ultrasonography. Kidney360. 2021 May 27;2(8):1326–1338.

9) Innes B, Levinson A, Abbasi A, et al. Venous excess ultrasound score (VEXUS) guided fluid management in septic shock: a Pilot trial. Critical Care Medicine. 2024 January 52(1):p S84.

10) Beaubien-Souligny W, Rola P, Haycock K, et al. Quantifying systemic congestion with Point-Of-Care ultrasound: development of the venous excess ultrasound grading system. Ultrasound J. 2020;12:16.

11) Soliman-Aboumarie H, Denault AY. How to assess systemic venous congestion with point of care ultrasound. Eur Heart J Cardiovasc Imaging. 2023;24:177–180.

12) Lang RM, Badano LP, Mor-Avi V, et al. Recommendations for cardiac chamber quantification by echocardiography in adults: an update from the American Society of Echocardiography and the European Association of Cardiovascular Imaging. J Am Soc Echocardiogr. 2015 Jan;28(1):1–39.e14.

13) Belletti A, Lerose CC, Zangrillo A, et al. Vasoactive-Inotropic Score: Evolution, Clinical Utility, and Pitfalls. J Cardiothorac Vasc Anesth. 2021 Oct;35(10):3067–3077.

14) Delgado C, Baweja M, Crews DC, et al. A Unifying Approach for GFR Estimation: Recommendations of the NKF-ASN Task Force on Reassessing the Inclusion of Race in Diagnosing Kidney Disease. Am J Kidney Dis. 2022 Feb;79(2):268–288.e1.

15) Guinot PG, Bahr PA, Andrei S, et al. Doppler study of portal vein and renal venous velocity predict the appropriate fluid response to diuretic in ICU: a prospective observational echocardiographic evaluation. Crit Care. 2022 Oct 5;26(1):305.

16) Longino A, Martin K, Leyba K, et al. Correlation between the VExUS score and right atrial pressure: a pilot prospective observational study. Crit Care. 2023 May 26;27(1):205.

17) Dimopoulos S, Antonopoulos M. Portal vein pulsatility: An important sonographic tool assessment of systemic congestion for critical ill patients. World J Cardiol. 2024 May 26;16(5):221–225.

18) Wong A, Olusanya O, Watchorn J, et al. Bramham K, Hutchings S. Utility of the Venous Excess Ultrasound (VEXUS) score to track dynamic change in volume status in patients undergoing fluid removal during haemodialysis - the ACUVEX study. Ultrasound J. 2024 Mar 27;16(1):23.

19) Ostermann M, Hall A, Crichton S. Low mean perfusion pressure is a risk factor for progression of acute kidney injury in critically ill patients - A retrospective analysis. BMC Nephrol. 2017 May 3;18(1):151.

20) El Nihum LI, Manian N, Arunachalam P, et al. Renal Dysfunction in Patients with Left Ventricular Assist Device. Methodist Debakey Cardiovasc J. 2022 Sep 6;18(4):19–26.

21) Roehm B, Hedayati S, Vest AR, et al. Long-Term Changes in Estimated Glomerular Filtration Rate in Left Ventricular Assist Device Recipients: A Longitudinal Joint Model Analysis. J Am Heart Assoc. 2023 Feb 7;12(3):e025993.

22) Tang WHW, Bakitas MA, Cheng XS, et al. Evaluation and Management of Kidney Dysfunction in Advanced Heart Failure: A Scientific Statement From the American Heart Association. Circulation. 2024 Oct 15;150(16):e280–e295.

23) Verstreken S, Beles M, Oeste CL, et al. eGFR slope as predictor of mortality in heart failure patients. ESC Heart Fail. 2025 Apr;12(2):1217–1226.

24) Manca P, Nuzzi V, Mulè M, et al. Gaps and Knowledge in the Contemporary Management of Acute Right Ventricular Failure. Circ Heart Fail. 2025 Apr 14:e012030.

25) Nonaka H, Lu LY, Bono NG, et al. Right heart failure after left ventricular assist device implantation: latest insights and knowledge gaps on mechanism and prediction. Front Cardiovasc Med. 2025 May 22;12:1586389.

26) Adamopoulos S, Bonios M, Ben Gal T, et al. Right heart failure with left ventricular assist devices: Preoperative, perioperative and postoperative management strategies. A clinical consensus statement of the Heart Failure Association (HFA) of the ESC. Eur J Heart Fail. 2024 Nov;26(11):2304–2322

27) Kan JY, Arishenkoff S, Wiskar K. Demystifying Volume Status: An Ultrasound-Guided Physiologic Framework. Chest. 2025 Jun;167(6):1667–1683.

28) Zucchetti O, Iacovoni A, Vittori C, et al. Longitudinal evaluation of congestion and renal function in advanced heart failure patients treated with a left ventricular assist device. Eur Heart J Suppl. 2022;24(Suppl_C):suac012.218.

29) Loungani RS, Mentz RJ, Agarwal R, et al. Biomarkers in Advanced Heart Failure: Implications for Managing Patients With Mechanical Circulatory Support and Cardiac Transplantation. Circ Heart Fail. 2020 Jul;13(7):e006840.

30) Longino AA, Martin KC, Leyba KR, et al. Reliability and reproducibility of the venous excess ultrasound (VExUS) score, a multi-site prospective study: validating a novel ultrasound technique for comprehensive assessment of venous congestion. Crit Care. 2024 Jun 11;28(1):197.

